# Are women underrepresented in cerebrovascular disease clinical trials? A systematic review using the FDA database

**DOI:** 10.1101/2020.11.09.20227207

**Authors:** Xiaobo Zhong, Qing Hao

## Abstract

**BACKGROUND AND PURPOSE:** Clinical trials provide essential evidence in the US Food and Drug Administration (FDA) drug approval process. Women were underrepresented in the enrollment of clinical trials for many medical conditions other than cerebrovascular disease (CVD). We investigated women’s participation in FDA-registered CVD-related interventional clinical trials and assessed whether women CVD patients in the United States were underrepresented. We also evaluated the association between the underrepresentation and the features of design and operation among these trials.

**METHODS:** We systematically reviewed the phase 2 and 3 CVD-related interventional trials started in 2002–2017 in the FDA database. The proportions of women enrolled in these trials and the proportion-to-prevalence ratios (PPRs), defined as the ratios of the proportions of women enrolled in these trials to the proportion of women prevalent among US CVD patients during the same period, were calculated and summarized by a meta-analysis approach. We used boosted regression tree, a machine learning model, to identify the determinants of women’s underrepresentation in CVD-related trials.

**RESULTS:** According to our selection criteria, we analyzed the data of 145 CVD-related trials, among which 40.9% (95% CI: 38.3–43.5%) of the patients enrolled were women. Their PPR was estimated to be 0.843 (95% CI: 0.796–0.890). We found that four factors substantially influenced women’s underrepresentation in these trials: the number of enrollment sites, the starting year, randomization, and academic institution sponsorship.

**CONCLUSIONS:** Generally, women with CVD in the United States were underrepresented in FDA registered trials started in 2002–2017. In addition, trials with greater numbers of enrollment sites, randomization, and non-academically sponsored trials had a higher risk of underrepresenting women with CVD in the United States. Investigators should take these factors into consideration in clinical trial design in the future, by either increasing women’s participation or stratifying the enrollment by gender.

## INTRODUCTION

Cerebrovascular disease (CVD), mainly including ischemic stroke, hemorrhagic stroke and subarachnoid hemorrhage, affects more than 7 million people each year and is one of the leading causes of disability and death in the United States.^1-2^ CVD can occur at any age, but it affects women and men differently.^3^ The gender differences in the epidemiology, symptomatology, and medical care of CVD patients have been well established.^4–9^ Compared to men, women generally have a higher mortality rate and worse clinical outcomes related to CVD due to their higher life expectancy, greater number of comorbidities, and more severe disease burden.^10^ Clinical trials provide essential evidence of the safety and effectiveness of drugs, which are viewed as pivotal in the US Food and Drug Administration (FDA) drug approval process.^11^ In practice, the results of gender-based analysis are not available in all clinical trial reports. It is more common to see that a clinical trial makes conclusions based on the average effect across women and men. However, in a clinical trial in which women were underrepresented, the proportion of women enrolled is lower than the proportion of women’s nationally prevalence among CVD patients. Therefore, the result could be biased toward men and lead to a suboptimal conclusion on the outcome of interest.^12^ Our study aimed to evaluate the proportion of women enrolled in FDA-registered CVD-related clinical trials started between 2002 and 2017 and assess whether women with CVD in the United States were underrepresented in these trials.

Historically, women were reported to be underrepresented in some clinical trials in the United States.^13,14^ To control for the potential impact of gender disparity, in 1993, the FDA started implementing guidelines for encouraging women to participate in clinical trials.^15^ A series of policies were later added to FDA regulations to strengthen the efforts to increase women’s participation.^16–21^ These actions gradually increased women’s involvement in some diseases, but their enrollment in CVD-related clinical trials has not been thoroughly studied. Burke and colleagues (2011) studied 22 phase 3 stroke trials sponsored by the National Institute of Neurological Disorders and Stroke (NINDS) between 1985 and 2008, in which 37.8% of the trial participants were women. The results suggested that women were likely underrepresented in these trials.^22^ On the other hand, several questions remain unclear in this field. First, other than the NINDS-funded trials, what has been the overall proportion of women participating in CVD-related trials in the United States, including those not funded by the NINDS, and has there been any improvement in more recent years, after 2008? Second, can women enrolled in CVD-related trials reasonably represent the prevalence of women with CVD in the United States? Third, if women were underrepresented in CVD-related trials, what were the possible design or operational features that could explain and predict the underrepresentation?

Motivated by these questions, using the FDA database, we conducted a systematic review of clinical trials that led to the approvals of CVD-related interventions and started between 2002 and 2017. We studied the temporary trends of women’s participation in these trials and obtained an overview of the underrepresentation of women by comparing the proportion of women enrolled in CVD trials with the proportion of women in the CVD patient population in the United States across the 16-year study period, using a meta-analysis approach. Information about a series of design and operational features were collected from these trials and analyzed using a boosted regression tree (BRT) model, a machine learning technique with an advantage in building a flexible predictive model for small-size datasets.^23^ The findings will benefit policymakers in guiding the enrollment of CVD-related clinical trials in the future.

## METHODS

### Study Design and Data Sources

The data used in this retrospective cohort study consisted of three parts that are all publicly available. The first part provided information about the enrollment of women and the design and operational feature variables of the CVD clinical trials. These data were obtained from the FDA database (https://clinicaltrials.gov). The second part of the study data contained information about the annual prevalence rates of CVD for women and men in the United States between 2002 and 2017, published in *Heart Disease and Stroke Statistics* of Circulation.^24^ The same definition of CVD were used in Parts 1 and 2. The third part of the data contained information on the size of the general US population by gender for every year during the study period, obtained from the US Census Bureau (www.census.gov/data). All the analyses were conducted based on trial-level data, and no individual-level data were included. The analytical dataset is available by the corresponding author (Dr. Xiaobo Zhong).

### Definition of Cerebrovascular Disease

We applied the definition of cerebrovascular disease as in *Heart Disease and Stroke Statistics* (i.e., ICD-9: 430–438; ICD-10: I60–I69), an annually updated statistical report by the American Heart Association.^24^ This report is generally viewed as the nationally representative source for monitoring CVD patients in the United States. Table I in the Supplementary Material lists all the conditions corresponding to these ICD-9/10 codes.

### Selection of Study Trials

We investigated women’s participation in clinical trials started in 2002–2017 that led to the FDA’s approvals of CVD-related interventions in the United States. The flowchart in Figure I of Supplementary Materials shows the details of selecting study trials from FDA database. The conditions included in the definition of cerebrovascular disease in the *Heart Disease and Stroke Statistics* (ICD-9: 430–438; ICD-10: I60–I69) were used when searching relevant trials in FDA database. A total of 2865 CVD-related interventional trials were identified and included in the selection procedure for building the analytical data. We excluded 1290 early phase trials, including 194 feasibility trials and 1096 phase 1 trials, due to small sample sizes and the primary goals of assessing safety and feasibility instead of effectiveness. We also excluded 341 phase 4 trials because these were post-market surveillances after FDA approvals. Trials that were suspended (N = 11), terminated (N = 187), withdrawn (N = 48), ongoing (N = 48), or of unknown completion status (N = 198) by the time of data collection were excluded, because they usually do not publish complete information of enrollments. Similarly, completed trials that did not report any results (N = 535) were excluded from the study data. We excluded trials without any enrollment site in the United States (N = 55), because these were typically non-US regional trials to provide data for local government agencies’ approval, instead of targeting the US market. We further excluded seven trials with sample sizes of less than 10 patients. A total of 145 clinical trials were finally selected for data analysis.

### Study Variables

The proportion of women enrolled in a trial was calculated as the number of women enrolled in the trial divided by the total number of patients enrolled in the trial. For each clinical trial selected, we collected information on a series of design and operational feature variables, including the started year of the trial, the number of enrollment sites, the phase (2 vs. 3), the trial design (randomized vs. non-randomized), and the sponsor type (the National Institutes of Health [NIH]/other US agency, an academic institution, or industry). A trial could be funded by multiple types of sponsors. For example, the Combined Approach to Lysis Utilizing Eptifibatide and Rt-PA in Acute Ischemic Stroke-Enhanced Regimen (CLEAR-ER) is a multi-center phase 2 trial sponsored by both the NIH and an academic institution (University of Cincinnati).^25^

### Statistical Analysis

We used statistical software R version 3.6.1 (R Foundation, Vienna, Austria) for data analysis.^26^ All the confidence intervals built in this study were two-sided at the confidence level of 95%. The study trials’ features were summarized by descriptive statistics, including the median (with an interquartile range [IQR]) for continuous variables and frequencies (with percentages) for categorical variables. We implemented a three-step approach to analyze the enrollment of women in CVD-related trials as follows.

### Proportion of Enrollment

In the first step, we conducted a meta-analysis to estimate the proportion of women enrolled in CVD trials across the study period of 2002–2017.^27^ Specifically, for each trial, we calculated the proportion of women who participated in it and constructed a weight based on the inverse of the variance of the estimated probability of a woman being enrolled in this trial.^28^ We then summarized the proportions of women participation across all the study trials into a single proportion with a 95% confidence interval using a random-effect meta-analysis model.^29^ A logarithmic transformation was applied to the calculation to avoid possible estimation bias due to the observed proportions with extremely high or low values.^30^ The meta-analysis model took into account the impact on the summarized proportion due to different sample sizes. A trial with a larger sample size was assigned more weight, thus having a greater impact on the summarized results. We also repeated the analysis and estimated the proportion of women participation within each subgroup of the feature variables. The random-effect meta-analysis models were built using the R package *metafor*.^31^

### Participation-to-prevalence Ratio (PPR)

In the second step, we assessed the representation of women in the FDA-registered CVD trials relative to the overall proportion of women among CVD patients in the United States. The PPR, defined as the ratio of the proportion of women enrolled in a trial to the proportion of women among US CVD patients in the United States, was calculated for each of the study trials as^32^

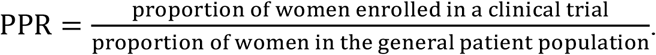

For example, the CLEAR-ER trial (NCT00894803) was designed to compare the efficacy of the combination therapy of rt-PA and eptifibatide versus rt-PA only for CVD patients. This trial was initiated in 2009 and enrolled 126 CVD patients, 60 of whom were women. The proportion of women enrolled in this trial was 0.476. Based on the prevalence rates of CVD and population sizes by gender in 2009, we calculated the proportion of women among the CVD patient population in 2009 to be 0.495 (Table II in Supplementary Material). Consequently, we calculated the PPR as 0.962 (= 0.476/0.495). Similar to the first step, the PPRs were then summarized into one proportion with a 95% confidence interval by a random-effect meta-analysis model overall and within each subgroup of interest. A PPR of one indicates that women were correctly represented in the CVD trial. Therefore, a summarized PRR with a 95% confidence interval of less than one suggests that women were underrepresented in the CVD clinical trials.

### Boosted Regression Tree (BRT) Model

In the third step, we fit the BRT model to identify the major determinants of women’s underrepresentation in CVD trials and quantify the associations between the identified determinants and the underrepresentation using the approach recommended by Elith (2008).^33^ The binary outcome of women’s underrepresentation in a clinical trial was defined as a PPR of less than one. The BRT model was formed to predict the probability of underrepresentation via the logit link function. The explanatory variables included the started year of the trial, the number of enrollment sites, the phase, the trial design, and whether the trial was funded by the NIH/other US agency, an academic institution, or industry. We used cross-validation to select the values of design parameters (i.e., bag fraction, tree complexity, learning rate, and the number of trees) for the optimal BRT model, defined as that with the smallest value of predictive deviance. We selected the explanatory variables included in the final BRT model based on their relative influences. Any variable with a relative influence of less than 5% was viewed as having a small impact on the women’s underrepresentation in CVD trials and was thus dropped from the final model. The effects of explanatory variables on the outcome were described using partial dependence plots. The BRT models were fitted by the R package *gbm*.^34^

## RESULTS

There were 145 CVD-related interventional clinical trials selected for analysis, including 80 (55%) phase 2 and 65 (45%) phase 3 trials (Table 1). Forty-five percent (N = 66) of the study trials started before 2010, and 55% (N = 79%) started after January 1, 2010. The median number of enrollment sites for these trials was eight, with 25th and 75th percentiles of 1 and 50, respectively. Forty-five trials (31%) enrolled patients at a single site. Among the multi-center trials, 80 (55%) trials had fewer than 100 sites, and 20 (14%) enrolled patients at ≥100 sites. Seventy percent of the trials were designed with randomization. These 145 trials enrolled a total of 251,965 patients.

**Table 1.**
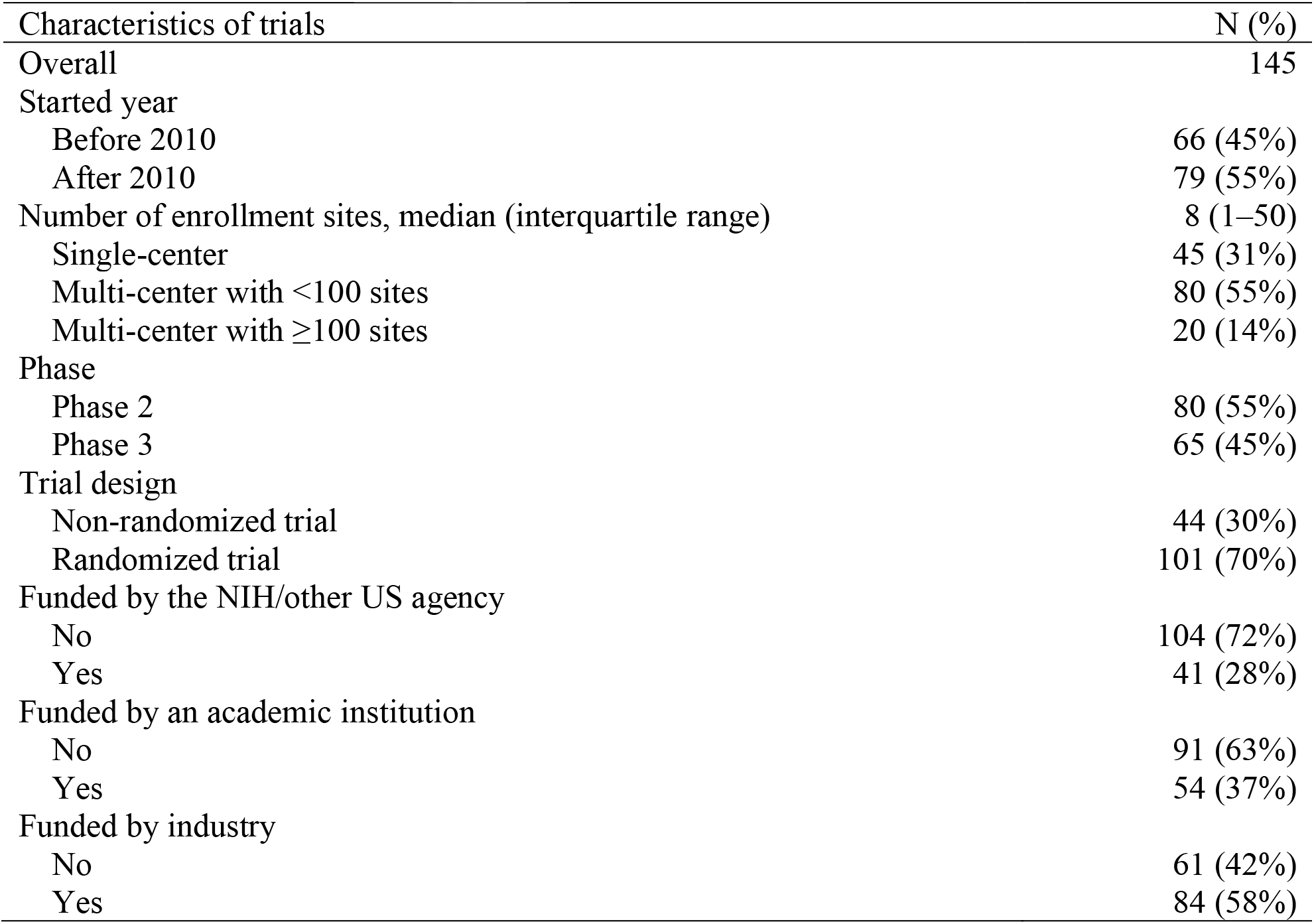
Characteristics of FDA-registered CVD-related clinical trials started in 2002-2017

| Characteristics of trials | N (%) |
| --- | --- |
| Overall | 145 |
| Started year |  |
| Before 2010 | 66 (45%) |
| After 2010 | 79 (55%) |
| Number of enrollment sites, median (interquartile range) | 8 (1–50) |
| Single-center | 45 (31%) |
| Multi-center with <100 sites | 80 (55%) |
| Multi-center with $\geq 100$ sites | 20 (14%) |
| Phase |  |
| Phase 2 | 80 (55%) |
| Phase 3 | 65 (45%) |
| Trial design |  |
| Non-randomized trial | 44 (30%) |
| Randomized trial | 101 (70%) |
| Funded by the NIH/other US agency |  |
| No | 104 (72%) |
| Yes | 41 (28%) |
| Funded by an academic institution |  |
| No | 91 (63%) |
| Yes | 54 (37%) |
| Funded by industry |  |
| No | 61 (42%) |
| Yes | 84 (58%) |

Figure 1 plots the proportions of women enrolled in the CVD trials. The solid line shows a decreasing trend in these proportions across the 16-year study period. The dashed line shows the trend of the proportion of women among general CVD patients in the United States during the same period. The proportions of women enrolled in the trials were consistently lower than those among US CVD patients. Table 2 provides the summarized proportions of women enrolled in CVD trials based on the meta-analysis model. The overall proportion of women enrolled in CVD trials was 0.409 (95% CI, 0.383, 0.435). The proportion was higher in phase 2 trials than in phase 3 (0.438 [0.403, 0.474)] vs. 0.381 [0.344, 0.419]). The proportion decreased as the number of sites increased, from 0.470 (0.432, 0.509) for single-center trials to 0.329 (0.283, 0.378) for multi-center trials with ≥100 sites. The proportions of women enrolled in trials funded by academic institutions, industry, and the NIH/other US agencies were 0.462 (0.421, 0.503), 0.396 (0.367, 0.426), and 0.386 (0.321, 0.454), respectively.

**Table 2.**
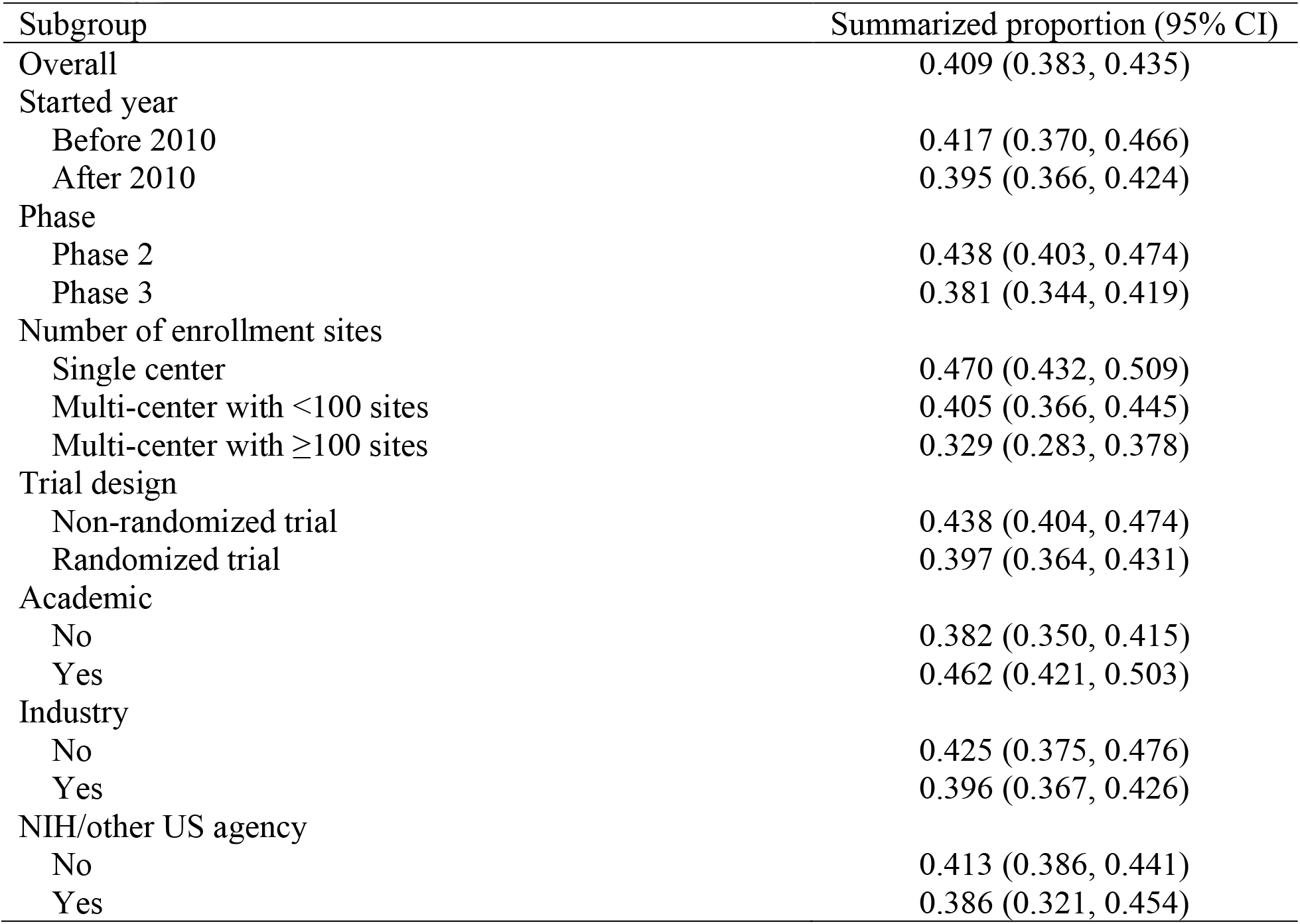
Proportions of women enrolled in FDA-registered CVD-related clinical trials from the meta-analysis

The overall PPR of women in CVD trials was 0.843 (0.796, 0.890), as shown in Figure 2. In the subgroup analysis, the PPR of trials funded by academic institutions was (0.938 [0.870, 1.007]), which was not significantly lower than one. In the meanwhile, the PPRs were 0.813 (0.753, 0.872) for industry-funded trials and 0.820 (0.725, 0.914) for trials funded by the NIH/other US agency (0.820 [0.725, 0.914]). The PPR decreased as the number of sites increased, from 0.942 (0.868, 1.106) for single-center trials to 0.844 (0.778, 0.910) for multi-center trials with fewer than 100 sites, and then to 0.649 [0.561, 0.736] for multi-center trials with ≥100 sites.

**Figure 1.**
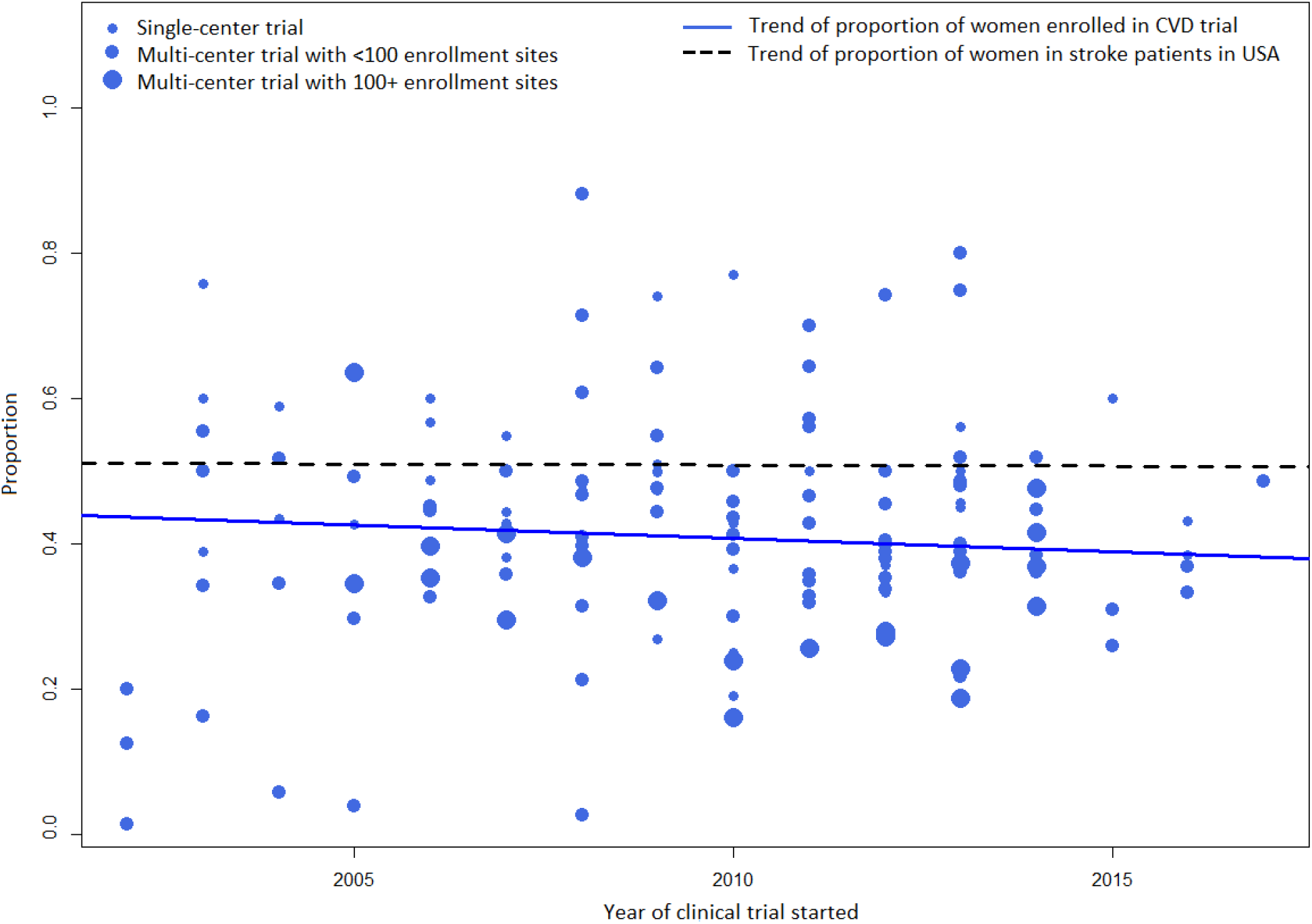
Proportions of women enrolled in FDA-registered interventional CVD-related clinical trials between 2002 and 2017

**Figure 2.**
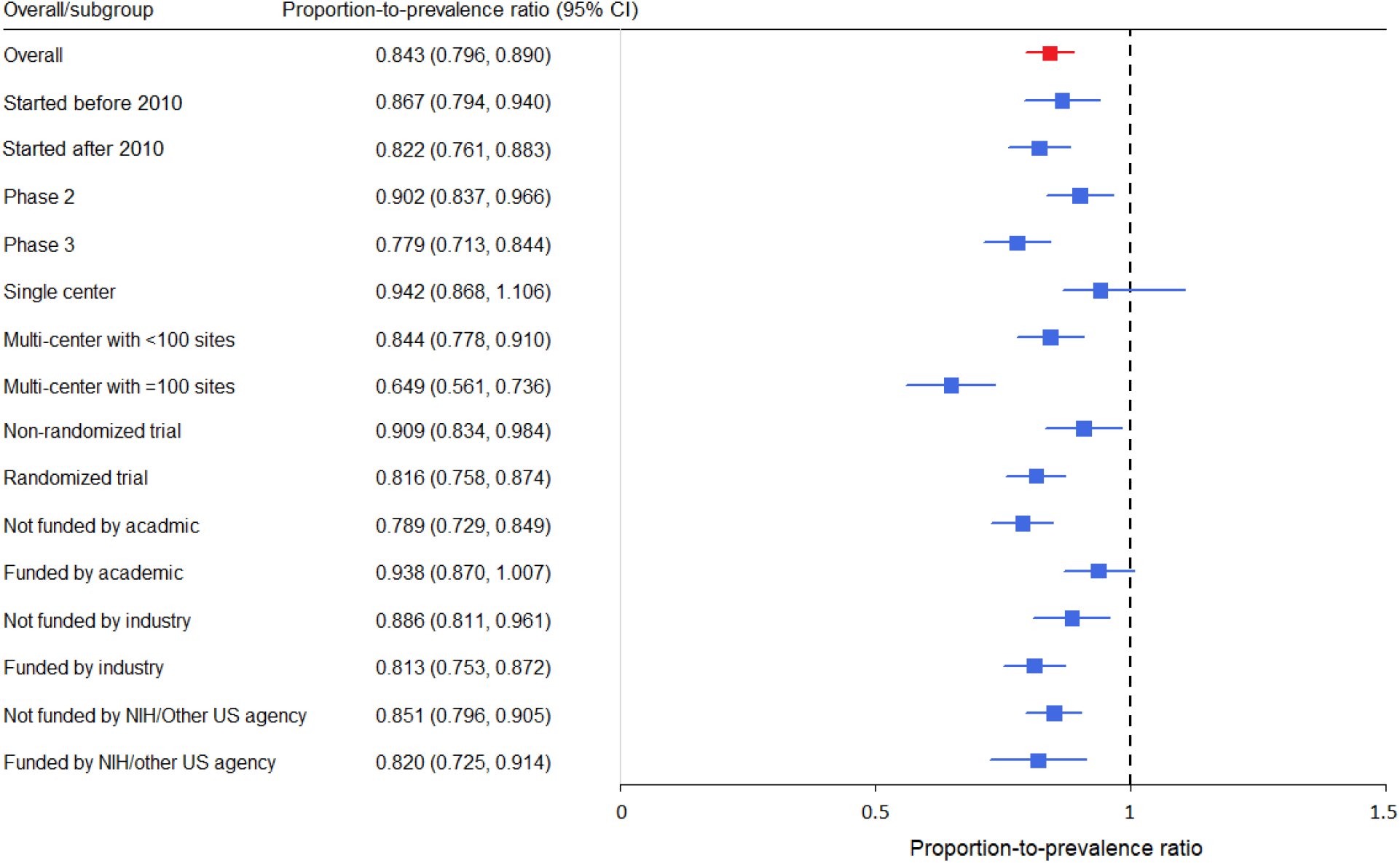
Proportion-to-prevalence ratios of women enrolled in FDA registered CVD-related clinical trials from the meta-analysis

We fitted a series of BRT models with five-fold cross-validation based on different combinations of design parameters. The optimal model was built based on a bag fraction of 0.5, a learning rate of 0.001, and a tree complexity of 5. The impacts of the explanatory variables on the outcome in the BRT model were quantified by relative influences. These relative influences were scaled so that their sum equaled one. A higher relative influence value indicates a greater impact. The first column in Table 3 provides the relative influences under the full BRT model, containing all eight explanatory variables. Sponsorship by the NIH/other US agencies or by industry, the phase, and the type of intervention had small relative influences (<5%) and were thus excluded from the variable selection. The final BRT model included four explanatory variables, which were the number of enrollment sites (relative influence 54.2%), the started year the trial (29.6%), the trial design (10.9%), and academic institution sponsorship (5.4%).

**Table 3.** Relative influences of the explanatory variables in the full and final boosted regression tree models

| Explanatory variables | Full model<br>(AUC = 0.824) | Final model<br>(AUC = 0.823) |
| --- | --- | --- |
| Number of enrollment sites | 49.6% | 54.3% |
| Started year | 27.1% | 29.1% |
| Trial design | 8.0% | 10.3% |
| Sponsored by an academic institution | 5.1% | 6.3% |
| Sponsored by the NIH/other US agency | 4.9% | - |
| Sponsored by industry | 2.7% | - |
| Phase | 2.6% | - |

\*AUC: Area under the receiver operating characteristic curve

Figure 3 shows four partial dependence plots, each corresponding to an explanatory variable in the final BRT model. The horizontal axis is the value of the explanatory variable. The vertical axis is the predicted odds of the underrepresentation of women, based on the final BRT model. The predicted values were standardized, and the result in each panel was adjusted for the three other covariates in the final model. Panel A shows that the predicted odds of the underrepresentation of women increased dramatically as the number of enrollment sites increased from about 20 to 100. Meanwhile, the predicted odds were relatively stable for those with more than 100 sites. In Panel B, adjusted for other covariates, the odds of the underrepresentation of women in CVD trials dropped from 2007 to 2009 and then increased until 2014. Panel C shows that randomized trials were more likely to underrepresent women than a non-randomized trial. Panel D shows that trials not funded by any academic institution were more likely to underrepresent women in CVD trials.

**Figure 3.**
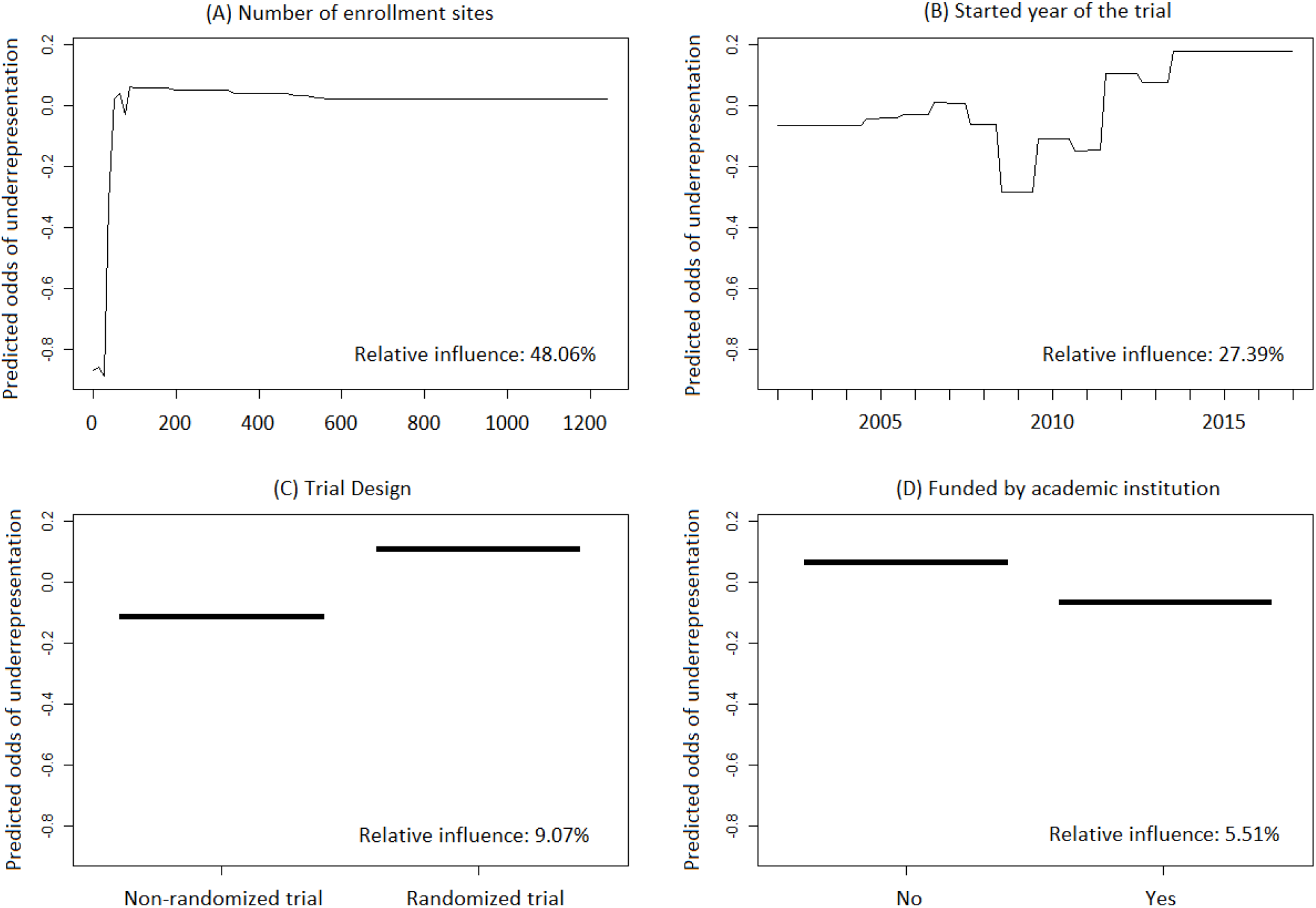
Partial dependence plots for the four explanatory variables in the final Boosted Regression Tree (BRT) model for predicting the underrepresentation of women in FDA-registered CVD-related clinical trials (Higher value of predicted odds indicates that women were more likely to be underrepresented in CVD trials.)

## DISCUSSION

The concept of gender in public health research differs from that of sex, which refers to the biological construct (e.g., reproductive organs and chromosomal complement) of male and female patients.^35^ Gender refers to the cultural, environmental, and psychosocial identification of being a man or woman. In practice, clinical trials typically report gender instead of sex. Therefore, our study focused on women CVD patients enrolled in clinical trials.

Concern about whether clinical trial enrollment correctly represents gender disparity in the real-world patient population has long existed.^19,20^ Tsivgoulis et al. (2017) demonstrated how the possible selection bias introduced by women’s underrepresentation will affect clinical trials’ conclusions.^12^ Although women’s participation in clinical trials has improved during the last two decades, women’s representation in clinical trials has still varied by disease indicators.^20,21^ For some diseases, such as cancer, heart disease, acute coronary syndrome, and osteoporosis, women were still underrepresented in clinical trials.^36–41^ By the time we initiated this project, two studies related to women enrolled in CVD-related clinical trials were conducted. Burk et al. (2011) reported the underrepresentation of women in 22 NINDS-funded CVD trials.^22^ In the other study, women were underrepresented, with less than 30% participants being women in a series of randomized clinical trials assessing antiplatelet agents for preventing CVD by 2012.^42^ Our work extended the research of women participation to all the CVD-related FDA registered interventional trials across a longer period (i.e., 2002–2017), and the findings were consistent with the two previous studies. Our study provides the most extensive observations (i.e., 145 trials) in studying women’s participation and representation in CVD-related trials.

The known factors that have deterred women from participating in clinical trials include difficulties in accessing enrollment sites, family responsibilities, cultural barriers, socioeconomic status, and concerns about negative outcomes.^43–45^ On the other hand, the associations between these factors and women’s underrepresentation have never been validated in the CVD patient population. In this study, we successfully identified four explanatory factors with substantial influences on the underrepresentation of women CVD patients. Among these factors, the number of enrollment sites had the highest impact on underrepresentation. Our model depicted the most critical range of enrollment sites as between 20 and 100, where the chance of underrepresentation dramatically increased as the number of sites increased. Our model also found that randomization was associated with higher odds of underrepresentation. Considering that almost all the clinical trials with more than 50 sites were randomized trials and non-randomized trials were typically small-size proof-of-concept trials, the findings for these two factors were consistent with each other. Similarly, trials sponsored by academic institutions were associated with low odds of women’s underrepresentation in CVD trials. These academic sponsored trials were usually of small sample size, with a relatively low number of sites. These findings need to be further validated in future studies and can help improve women’s underrepresentation at the design stage of a CVD trial in the future.

An advantage of our study is its rigorous statistical approach, including meta-analysis and machine learning models, for inference. We applied a random-effect model in the meta-analysis to synthesize the proportions of women enrolled in CVD trials and the PPRs. By this method, we constructed a weight that took into account the influence due to the different sample sizes of CVD trials. This weight helped minimize the within-trial variations of the estimated proportion and PPR. Meanwhile, the random-effect model assumed variation in the proportions of women enrolled (or their PPR) across different trials and thus helped minimize between-trial variation.^46^

We used the BRT model to identify the driving factors of the underrepresentation of women in CVD trials. This machine learning model has two advantages compared to traditional statistical models, such as the generalized linear regression model (GLM) and the generalized additive model (GAM).^47^ First, it generally has better prediction performance. Second, it has unique flexibility in describing the association between a continuous explanatory factor and the outcome.^48^ A traditional GLM or GAM model typically assumes a linear, quadratic or cubic relationship between a continuous predictor and the binary outcome or translates it into a categorical variable according to some cutoff value. In a BRT model, with the help of a large number of trees, we can depict the relation between a continuous predictor and the outcome in a flexible manner, as we see in panels A and B of Figure 3.

A limitation of this study is that the analysis was based on a complete case analysis. On the other hand, trials that did not report enrollment information due to suspension, termination, or withdrawal were excluded from the analysis. In other words, our inference procedure assumed missing completely at random. Ideally, the study should have included all the FDA registered interventional trials for CVD patients. However, in practice, few clinical trials that were suspended, terminated, or withdrawn would release enrollment information to the public.

Similarly, it is difficult to find any enrollment information for an ongoing trial. Therefore, we could not collect the data of these studies in a systematic review. Our inference assumed that the suspension, termination, or withdrawal of CVD-related trials was independent of women’s enrollment in these trials. That is to say, no investigator would decide to suspend, terminate, or withdraw a CVD trial because (or partially because) the trial failed to enroll a certain proportion of women. A combined dataset based on multiple accessible randomly selected suspended, terminated, and withdrawn clinical trials would be a possible solution to address this issue in the future.

## CONCLUSION

Generally, women with CVD in the United States were underrepresented in FDA-registered interventional clinical trials started between 2002 and 2017. This underrepresentation has been worsened in recent year. Trials with larger numbers of enrollment sites, randomized trials, and non-academically sponsored trials had a higher risk of underrepresenting women with CVD in the United States. Investigators should pay attention to these factors at the design stage of future CVD-related trials, by either increasing women’s participation or stratifying the enrollment by gender.

## Data Availability

The study data were obtained from FDA database.

https://www.clinicaltrials.gov/

