## Supplemental Table I for "Are women underrepresented in cerebrovascular disease clinical trials? A systematic review using the FDA database"

Supplementary Material

Table I. Diseases and conditions under the diagnosis codes for CVD

| Diagnosis code | Classification of disease |
|---|---|
| ICD-9: 430 | Subarachnoid hemorrhage |
| ICD-9: 431 | Intracerebral hemorrhage |
| ICD-9: 432 | Other and unspecified intracranial hemorrhage |
| ICD-9: 433 | Occlusion and stenosis of precerebral arteries |
| ICD-9: 434 | Occlusion of cerebral arteries |
| ICD-9: 435 | Transient cerebral ischemia |
| ICD-9: 436 | Acute, but ill-defined, cerebrovascular disease |
| ICD-9: 437 | Other and ill-defined cerebrovascular disease |
| ICD-9: 438 | Late effects of cerebrovascular disease |
| ICD-10: I60 | Nontraumatic subarachnoid hemorrhage |
| ICD-10: I61 | Nontraumatic intracerebral hemorrhage |
| ICD-10: I62 | Other and unspecified nontraumatic intracranial hemorrhage |
| ICD-10: I63 | Cerebral infarction |
| ICD-10: I65 | Occlusion and stenosis of precerebral arteries, not resulting in cerebral infarction |
| ICD-10: I66 | Occlusion and stenosis of cerebral arteries, not resulting in cerebral infarction |
| ICD-10: I67 | Other cerebrovascular diseases |
| ICD-10: I68 | Cerebrovascular disorders in diseases classified elsewhere |
| ICD-10: I69 | Sequelae of cerebrovascular disease |

The ICD-9 and ICD-10 codes for CVD were based on the *Heart Disease and Stroke Statistics* annual report.
