## Supplementary Table II for "Are women underrepresented in cerebrovascular disease clinical trials? A systematic review using the FDA database"

Supplementary Material

Table II. Prevalence of CVD among patients of age≥18 years by gender (females vs. males) in the United States between 2002 and 2017

| Year | Female | | | Male | | | Overall | | | Ratio of women to men among CVD patients |
|---|---|---|---|---|---|---|---|---|---|---|
| | Population size[*] | Prevalence rate[†] | Estimated N of patients[‡] | Population size[1] | Prevalence rate[2] | Estimated N of patients | Population size | Prevalence rate | Estimate N of patients | |
| 2002 | 108712 | 2.5% | 2717.80 | 100610 | 2.5% | 2515.25 | 209322 | 2.5% | 5233.05 | 1.08:1 |
| 2003 | 110190 | 2.5% | 2754.75 | 102431 | 2.5% | 2560.78 | 212621 | 2.5% | 5315.53 | 1.08:1 |
| 2004 | 110883 | 2.5% | 2772.08 | 103641 | 2.5% | 2591.03 | 214524 | 2.5% | 5363.10 | 1.07:1 |
| 2005 | 112409 | 2.6% | 2922.63 | 104966 | 2.5% | 2624.15 | 217375 | 2.6% | 5546.78 | 1.11:1 |
| 2006 | 113503 | 2.6% | 2951.08 | 106346 | 2.5% | 2658.65 | 219849 | 2.6% | 5609.73 | 1.11:1 |
| 2007 | 114880 | 2.8% | 3216.64 | 107843 | 2.6% | 2803.92 | 222723 | 2.7% | 6020.56 | 1.15:1 |
| 2008 | 115842 | 2.5% | 2896.05 | 108863 | 2.7% | 2939.30 | 224705 | 2.6% | 5835.35 | 0.99:1 |
| 2009 | 116946 | 2.5% | 2923.65 | 110027 | 2.7% | 2970.73 | 226973 | 2.6% | 5894.38 | 0.98:1 |
| 2010 | 118079 | 2.5% | 2951.98 | 111163 | 2.7% | 3001.40 | 229242 | 2.6% | 5953.38 | 0.98:1 |
| 2011 | 118892 | 2.5% | 2972.30 | 112301 | 2.7% | 3032.13 | 231193 | 2.6% | 6004.43 | 0.98:1 |
| 2012 | 121442 | 2.6% | 3157.49 | 113279 | 2.6% | 2945.25 | 234721 | 2.6% | 6102.75 | 1.07:1 |
| 2013 | 122481 | 2.6% | 3184.51 | 114448 | 2.7% | 3090.10 | 236929 | 2.6% | 6274.60 | 1.03:1 |
| 2014 | 129959 | 2.9% | 3768.81 | 115880 | 2.9% | 3360.52 | 245839 | 2.9% | 7129.33 | 1.07:1 |
| 2015 | 125196 | 2.7% | 3380.29 | 117051 | 2.7% | 3160.38 | 242247 | 2.7% | 6540.67 | 1.07:1 |
| 2016 | 126338 | 2.7% | 3411.13 | 118468 | 2.7% | 3198.64 | 244806 | 2.7% | 6609.76 | 1.07:1 |
| 2017 | 127156 | 2.8% | 3560.37 | 119168 | 2.8% | 3336.70 | 246324 | 2.8% | 6897.07 | 1.07:1 |
| 2018 | 128488 | 2.7% | 3469.18 | 120708 | 2.7% | 3259.12 | 249196 | 2.7% | 6728.29 | 1.06:1 |
| 2019 | 129262 | 2.8% | 3619.34 | 121300 | 2.9% | 3517.70 | 250562 | 2.8% | 7137.04 | 1.03:1 |

[*]The population sizes (numbers in 1000s) were obtained from the data released by the US Census Bureau (https://www.census.gov).

[†]The prevalence rates were based on the *Heart Disease and Stroke Statistics* annually updated report from *Circulation*.

[‡]The estimated number N of patients was calculated as the population multiplied by the prevalence rate.
