## Supplementary figures and images for "Are women underrepresented in cerebrovascular disease clinical trials? A systematic review using the FDA database"

### Supplementary Figure I

Supplementary Material

Figure I: Selection of study trials in FDA database

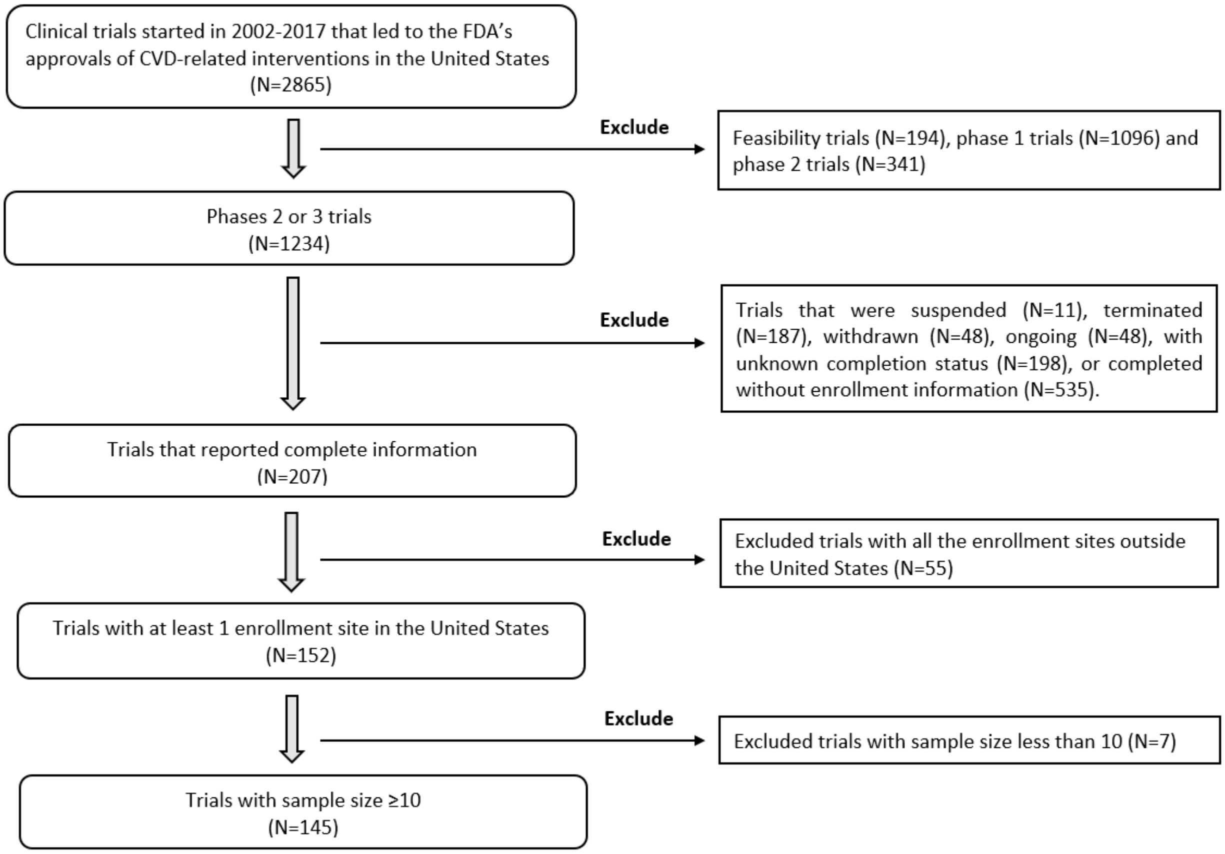
